# Prevalence and Clinical Correlates of Coronary Perivascular Inflammation in an Asymptomatic Adult Primary Prevention Population: An Assessment of the CT-Derived Fat Attenuation Index Score (FAI-Score)

**DOI:** 10.64898/2026.10.19.26364786

**Authors:** Diana Stan, Jonathan Fu, Monica Ngov, Michael Doney

## Abstract

**Background:** Coronary perivascular fat attenuation index score (FAI-Score) on coronary computed tomography angiography (CCTA) quantifies vascular inflammation and predicts cardiac mortality in symptomatic cohorts. However, its prevalence, anatomical distribution, and clinical correlates in asymptomatic primary prevention populations remain uncharacterized.

**Methods:** In an IRB-approved prospective study across two clinical sites, 84 consecutive asymptomatic adults (mean age 53.7 ± 8.6 years; 63.1% men) with no prior coronary artery disease underwent screening CCTA and comprehensive clinical, metabolic, and biomarker profiling. AI-standardized FAI-Score was quantified across three epicardial vessels (LAD, LCx, RCA). High and very high perivascular inflammation were pre-specified as any vessel ≥75th and ≥90th percentiles, respectively. Luminal stenosis and plaque were graded using CAD-RADS 2.0.

**Results:** Elevated FAI-Score (≥75th percentile) was present in 42.9% of participants (36/84; 95% CI 32.8% - 53.5%), with 17.9% (15/84; 95% CI 11.1% - 27.4%) exhibiting very high inflammation (≥90th percentile). Multi-vessel inflammation was identified in 17.9% (15/84), with the RCA most frequently involved (≥90th percentile in 13.1%). Standard circulating biomarkers, including ApoB, non-HDL-C, HbA1c, and PREVENT-ASCVD 10-year risk showed no association with binary inflammation. Systemic hs-CRP (median 0.8 mg/L) exhibited an inverse correlation with maximum vessel FAI-Score (Spearman ρ=−0.27, p=0.014). Men demonstrated higher rates of elevated FAI-Score than women (52.8% vs. 25.8%, p=0.016; ≥90th percentile: 26.4% vs. 3.2%, p=0.007). Strikingly, 46.2% (30/65) of participants with normal luminal anatomy (CAD-RADS 0) and 47.2% (25/53) with zero plaque burden harbored elevated FAI-Score; all 15 individuals with very high FAI-Score had CAD-RADS 0 (Fisher’s exact p=0.018). Body mass index was the sole secondary correlate surviving false discovery rate correction (AUC = 0.71, p=0.001, all-test FDR q=0.028). Regularized multivariable models showed modest out-of-sample discrimination for FAI-Score elevation (cross-validated AUC = 0.71).

**Conclusions:** Coronary perivascular inflammation is common among asymptomatic adults and frequently uncouples from structural plaque and luminal stenosis. Routine circulating biomarkers and clinical risk calculators fail to identify inflamed vessels, highlighting FAI-Score as an independent dimension of primary cardiovascular risk assessment.

## Introduction

Cardiovascular disease (CVD) remains the leading cause of death in the United States and worldwide, accounting for approximately 941,000 deaths in the United States in 2022 and an estimated 19.4 million deaths globally in 2021.[1,2] Coronary heart disease (CHD) was responsible for nearly 40% of US cardiovascular deaths, causing 371,506 deaths in 2022.[1] Despite substantial advances in prevention, diagnosis, and treatment, an estimated 800,000 Americans experience a new or recurrent myocardial infarction annually.[1] Estimated direct costs of CVD in the United States increased from $189.7 billion in 2012–2013 to $233.3 billion in 2020–2021.[1] Notably, sudden cardiac death can be the first recognized manifestation of coronary artery disease (CAD), underscoring the limitations of relying on symptom-triggered evaluation and the importance of identifying at-risk, asymptomatic individuals before a first major cardiovascular event.[3]

Decades of epidemiologic research have established a well-characterized set of modifiable risk factors for CAD. The landmark INTERHEART study demonstrated that nine conventional risk factors—including dyslipidemia, smoking, hypertension, diabetes, abdominal obesity, psychosocial factors, diet, physical activity, and alcohol consumption—collectively accounted for approximately 90% of the population-attributable risk for a first myocardial infarction across diverse global populations.[4] Several conventional risk factors are incorporated into contemporary prediction tools, including SCORE2 and the American Heart Association’s Predicting Risk of Cardiovascular Disease EVENTs (PREVENT) equations.[5,6] These tools inform primary prevention, with the 2026 ACC/AHA dyslipidemia guideline recommending PREVENT-ASCVD to guide risk-based lipid-lowering decisions.[7]

However, clinical risk estimates do not fully capture the presence and extent of subclinical atherosclerosis. In PESA (Progression of Early Subclinical Atherosclerosis), vascular ultrasound and coronary artery calcium (CAC) scoring identified subclinical disease in 58% of asymptomatic, middle-aged participants classified as having low 10-year Framingham risk.[8] More recently, REACT used carotid and femoral ultrasonography together with coronary computed tomography angiography (CCTA) to identify silent atherosclerosis across the assessed arterial territories in 57.1% of 16,808 adults aged 18–70 years without known atherosclerotic cardiovascular disease.[9] The Multi-Ethnic Study of Atherosclerosis (MESA) demonstrated that CAC scoring provides prognostic information beyond conventional risk factors, with a CAC score of zero supporting downward risk reclassification.[10,11] Nevertheless, the absence of coronary calcium does not exclude noncalcified plaque: the Miami Heart Study identified coronary plaque in 16% of asymptomatic participants with CAC scores of zero.[12] Together, these findings highlight the complementary information provided by vascular imaging and motivate investigation of imaging biomarkers that characterize coronary disease biology beyond plaque presence and extent.

Atherosclerosis is a dynamic process in which lipid accumulation and inflammation interact to promote plaque development and complications.[13] CANTOS provided randomized evidence that targeting inflammation can reduce cardiovascular events: among patients with prior myocardial infarction and elevated high-sensitivity C-reactive protein (hs-CRP), canakinumab 150 mg every three months reduced the primary cardiovascular endpoint by 15% relative to placebo without lowering LDL cholesterol (HR, 0.85; 95% CI, 0.74–0.98).[14] A pooled analysis of 31,245 statin-treated participants in PROMINENT, REDUCE-IT, and STRENGTH also found that hs-CRP was more strongly associated with subsequent cardiovascular events and mortality than LDL cholesterol.[15] These findings support residual inflammatory risk as a distinct, actionable pathophysiological domain in secondary prevention and motivate the development of imaging tools capable of assessing localized coronary vascular inflammation, complementing systemic markers such as hs-CRP.

CCTA permits indirect assessment of coronary inflammation through changes in adjacent perivascular adipose tissue. Antonopoulos and colleagues demonstrated that vascular inflammatory signaling inhibits adipocyte differentiation and lipid accumulation, shifting perivascular fat attenuation toward higher, less negative Hounsfield units. This response can be characterized using the Fat Attenuation Index (FAI).[16] In CRISP-CT, elevated right-coronary FAI was independently associated with cardiac mortality; a threshold of ≥−70.1 HU was associated with adjusted hazard ratios of 9.04 and 5.62 in the derivation and validation cohorts, respectively.[17] A subsequent meta-analysis found an association between elevated pericoronary fat attenuation and major adverse cardiovascular events (pooled HR, 3.29; 95% CI, 1.88–5.76), although between-study heterogeneity was substantial (I² = 75%).[18]

More recently, the Oxford Risk Factors and Non-invasive Imaging (ORFAN) study extended this evidence to standardized FAI-Score. Among 40,091 patients undergoing clinically indicated CCTA, a nested cohort of 3,393 patients with median follow-up of 7.7 years underwent evaluation of the prognostic value of FAI-Score and an integrated AI-Risk algorithm. Higher FAI-Score was independently associated with cardiac mortality and major adverse cardiovascular events, including among patients without obstructive CAD. The integrated algorithm, combining FAI-Score with clinical risk factors and coronary plaque information, also improved risk stratification.[19] Collectively, these data support coronary inflammation assessment as a distinct dimension of cardiovascular risk evaluation that provides prognostic information beyond conventional clinical and anatomical measures.

The major prognostic studies evaluated patients referred for clinically indicated CCTA.[17,19] Exploratory work has also assessed FAI-Score in selected asymptomatic populations, including a specialist lipid-clinic cohort.[20] Evidence remains limited, however, in asymptomatic adults undergoing comprehensive preventive evaluation. Characterizing the frequency, vessel distribution, and clinical correlates of elevated FAI-Score in this setting is an important step toward evaluating its potential role in preventive risk assessment.

The objectives of this cross-sectional study were to:

1. Characterize the prevalence and vessel-specific distribution of elevated FAI-Score in asymptomatic adults undergoing preventive CCTA at a dedicated preventive diagnostics center;
2. Examine associations with atherogenic lipoproteins, glycemic indices, hs-CRP, and PREVENT-ASCVD risk, and explore the ability of clinical and biochemical variables to classify person-level FAI-Score elevation using internally validated models; and
3. Examine relationships between FAI-Score elevation, coronary plaque presence, and luminal stenosis.

## Materials and Methods

### Study Design, Setting, and Ethical Oversight

This study was an investigator-initiated, prospective observational cohort study conducted across two clinical sites operated by Biograph International Inc., a preventive diagnostics and longevity healthcare organization in the United States. The study protocol was reviewed and approved by an independent Institutional Review Board (WCG, IRB Protocol No.: 20253603; IRB Study No.: 1399242). All enrolled participants provided written, prospective informed consent prior to undergoing imaging and laboratory evaluations.

### Study Population and Eligibility Criteria

The study cohort comprised a convenience sample of consecutive, self-referred asymptomatic adults aged 40 to 80 years presenting for comprehensive elective preventive cardiovascular screening, which included contrast-enhanced coronary computed tomography angiography (CCTA).

To isolate an authentic primary prevention cohort free of overt or occult clinical coronary artery disease, strict eligibility criteria were applied:

- **Inclusion Criteria:** (1) Age 40 to 80 years at the time of screening; (2) undergoing contrast-enhanced CCTA as part of a routine, comprehensive preventive health evaluation; (3) complete absence of history of angina, anginal equivalents, or exertional chest discomfort; (4) no documented history of coronary artery disease; and (5) capacity and willingness to provide informed consent.
- **Exclusion Criteria:** (1) Prior history of acute coronary syndrome (ST-segment elevation or non–ST-segment elevation myocardial infarction, unstable angina); (2) prior coronary revascularization (percutaneous coronary intervention [PCI] or coronary artery bypass graft [CABG] surgery); (3) current suspected symptoms of angina or anginal equivalents; (4) active or recent use of biologic anti-inflammatory therapeutics, systemic corticosteroids, or colchicine within the preceding 3 months, or cumulative lifetime exposure exceeding 6 months; (5) significant motion, calcification, or beam-hardening artifacts on CCTA precluding diagnostic interpretation; (6) CCTA examinations deemed non-diagnostic or lacking sufficient perivascular adipose tissue visualization for attenuation analysis; and (7) current pregnancy.

An analytical study cohort of 84 consecutive participants with complete, three-vessel perivascular fat attenuation profiling was finalized for primary analysis.

### Coronary CTA Acquisition, Plaque Grading, and Perivascular Adipose Tissue Analysis

All contrast-enhanced CCTA examinations were performed in accordance with Society of Cardiovascular Computed Tomography (SCCT) guidelines on contemporary multidetector CT scanners using prospective electrocardiographic gating, target heart rate control via pre-scan beta-blockade when indicated, and sublingual nitroglycerin administration. Examinations were interpreted by board-certified cardiovascular radiologists and cardiologists.

Structural plaque burden and luminal stenosis were graded according to the standardized Coronary Artery Disease - Reporting and Data System (CAD-RADS 2.0) guidelines. [21] Plaque characterization (presence, anatomical location, calcified versus non-calcified/soft components, and segment involvement) was quantified utilizing the CaRi-Plaque module (Caristo Diagnostics, Oxford, UK).

Digital Imaging and Communications in Medicine (DICOM) image datasets were securely transferred via encrypted cloud transmission to Caristo Diagnostics for perivascular fat attenuation analysis using the CaRi-Heart platform. The methodology of perivascular Fat Attenuation Index (FAI) and artificial intelligence-standardized FAI-Score calculation has been previously validated in large prospective cohorts. [16, 17, 19] In brief, technical scan parameters (e.g. tube voltage), anatomical factors related with the fat distribution around the arteries and basic demographics (age, sex) were standardized using an artificial intelligence algorithm to generate the vessel-specific FAI-Score, expressed as age- and sex-standardized percentiles. [22]

Person-level inflammation was defined a priori using Caristo clinical percentile cut-points:

- **Elevated/High Inflammation:** Any major epicardial vessel (LAD, LCx, or RCA) at or above the 75th percentile representing high inflammation.
- **Very High Inflammation:** Any major epicardial vessel at or above the 90th percentile.
- **Inflamed-Vessel Count:** The total number of inflamed epicardial vessels per participant (range: 0 to 3).
- **Max Vessel Percentile:** The highest percentile recorded across the three interrogated vessels within an individual.

### Clinical, Metabolic, and Circulating Biomarker Profiling

All participants underwent paired clinical, anthropometric, and laboratory profiling performed under fasting conditions concurrently with imaging:

**Circulating Lipids and Apolipoproteins:** Total cholesterol, high-density lipoprotein cholesterol (HDL-C), low-density lipoprotein cholesterol (LDL-C), triglycerides, non-HDL-C, apolipoprotein B (ApoB), and lipoprotein(a) [Lp(a)].
**Vascular and Systemic Inflammatory Markers:** High-sensitivity C-reactive protein (hs-CRP), total homocysteine, and oxidized LDL (oxLDL).
**Glycemic, Metabolic, and Fatty Acid Biomarkers:** Fasting plasma glucose, fasting serum insulin, hemoglobin A1c (HbA1c), the calculated Homeostatic Model Assessment of Insulin Resistance (HOMA-IR), and the OmegaQuant erythrocyte Omega-3 Fatty Acid Index.
**Multimodal Phenotyping:** Whole-body dual-energy x-ray absorptiometry (DEXA) for quantitative visceral adipose tissue (VAT) mass and volume; brain magnetic resonance imaging (MRI) for quantitative cerebral white matter hyperintensity (WMH) volume; and cardiopulmonary exercise testing (CPET) for absolute and body mass-indexed maximum oxygen uptake (VO2max).
**Cardiovascular Risk Algorithms:** 10-year atherosclerotic cardiovascular disease (ASCVD) risk was calculated using the 2024 American Heart Association PREVENT-ASCVD (Predicting Risk of cardiovascular disease EVENTs) equations [6].
**Clinical Covariates:** Standardized health history questionnaires extracted from the Biograph clinical electronic database captured baseline demographic parameters, smoking status (never, former, active), physician-diagnosed hypertension or type 2 diabetes, and active pharmacotherapies (antihypertensive agents, statins, GLP-1 receptor agonists). All data were anonymized and stored within an encrypted, audit-trailed data warehouse.

### Statistical analysis

Continuous variables are presented as either mean with standard deviation or median with interquartile range when absolute sample skewness was greater than 1. Categorical variables are presented as frequencies or percentages with Wilson score 95% confidence intervals without continuity correction calculated for key prevalence estimates. Outcomes for perivascular coronary inflammation were defined as having at least one vessel with a Fat Attenuation Index percentile of 75 or higher (“High Inflammation”) or 90 or higher (“Very High Inflammation”). Associations between continuous predictors and binary inflammation outcomes were evaluated via the Mann-Whitney U test using rank-biserial correlation for effect size. The binary inflammation and binary very high inflammation outcomes were defined as having at least one vessel at the 75th percentile or higher or 90th percentile or higher respectively for inflammation. Receiver-operating characteristic curves (ROC) and area under the ROC curves (AUC) were computed to evaluate the effectiveness of variables in predicting coronary inflammation outcomes. Spearman correlation coefficient, ρ, and Pearson correlation coefficient, r, were computed for rank and continuous variables, respectively.

Associations between categorical predictors and binary inflammation outcomes were evaluated by Pearson’s chi-squared test or Fisher’s exact test in the cases where expected cell count was fewer than five. Effect sizes were reported as conditional odds ratios with 95% confidence intervals.

Continuous inflammation outcomes included the Fat Attenuation Index (FAI-Score), defined as the maximum percentile across all three primary coronary vessels and each individual vessel’s percentile. Spearman rank correlations were calculated to evaluate associations with continuous biomarkers. Adjustments made to retain zero values included natural-log transforming hs-CRP, Lp(a), and oxLDL, and log(1 + CAC) for Coronary Artery Calcium score.

Multivariable Ordinary Least Squares (OLS) models with HC3 heteroskedasticity-robust standard error having inputs including ApoB, log-transformed hs-CRP, HbA1c, age, sex, and BMI predicted FAI-Score and individual vessel percentiles. The model output regression coefficient, 95 % confidence intervals, p-values, adjusted R², and variance inflation factors.

Benjamini-Hochberg False Discovery Rate (FDR) corrections were applied to univariate association tests: to Mann-Whitney U p-values for continuous predictors versus high and very high inflammation outcomes, to Fisher’s and chi-squared p-values for categorical predictors versus high and very high inflammation outcomes, and to Spearman p-values for FAI-Score and percentiles by vessel versus continuous FAI-Score and individual vessel percentiles.

In post hoc exploratory analysis, the relationship between sex and inflammation was explored before and after adjustment for BMI, by specifying models predicting inflammation by sex and sex as well as BMI. Post hoc analysis was also done to explore the weak inverse relationship between elevated hs-CRP and FAI-Score.

Analysis was performed in Python using pandas, NumPy, SciPy and statsmodels. Tables and figures were created in Python using Matplotlib and TableOne. Data was collated using Google CloudBigQuery Google Sheets and Google AppSheets.

## Results

### Study Population and Baseline Characteristics

The analytical cohort included 84 consecutive asymptomatic adults with complete three-vessel FAI-Score analysis, including 53 men (63.1%) and 31 women (36.9%). Baseline demographic and clinical characteristics are summarized in Table 1 and Table 2.

**Table 1.**
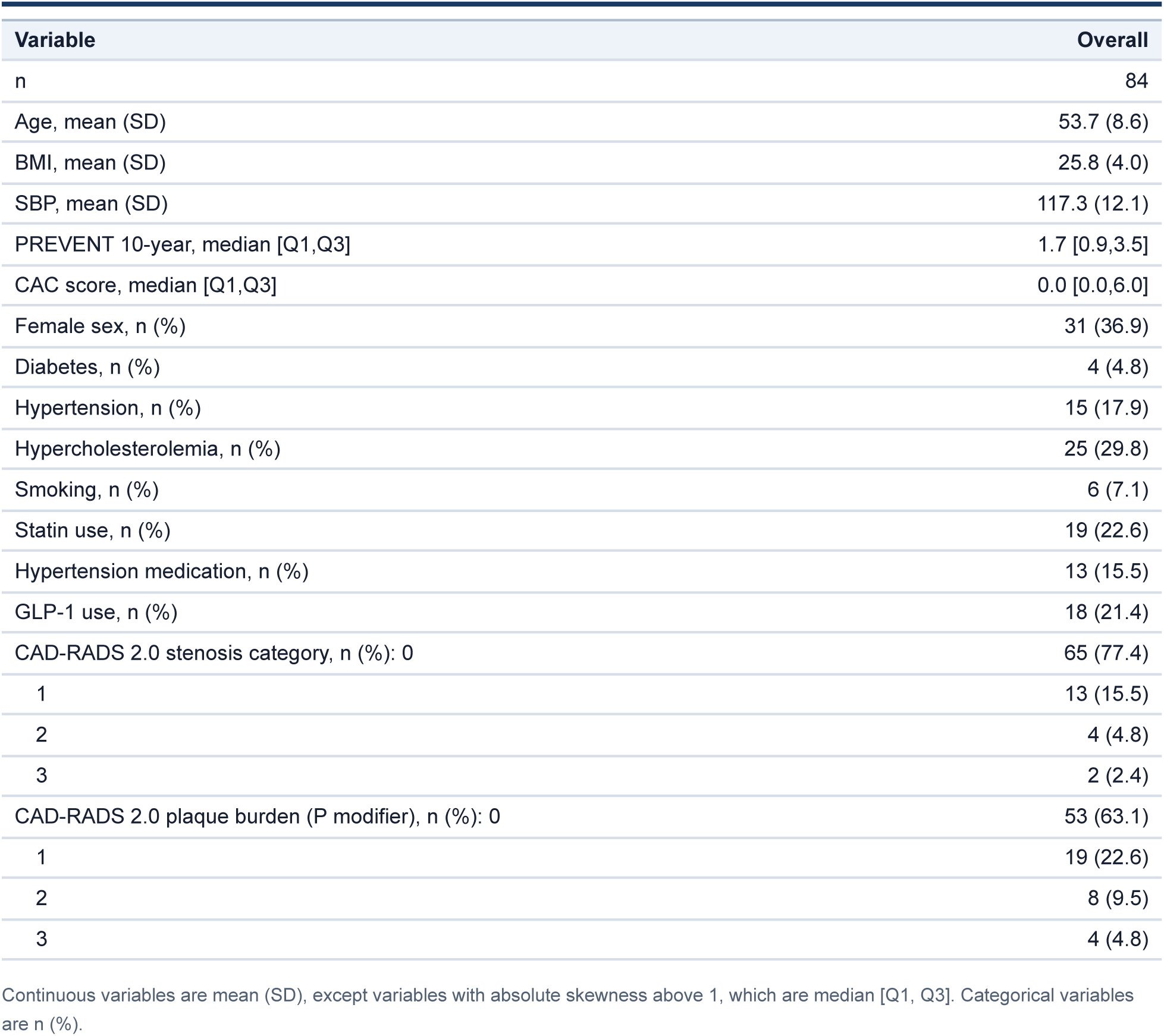
Cohort demographics.

**Table 2.** Clinical variables selected for association testing.

| Variable | Missing | Overall |
| --- | --- | --- |
| n |  | 84 |
| Non-HDL-C, mean (SD) | 0 | 136.9 (42.0) |
| HDL-C, mean (SD) | 0 | 65.8 (17.8) |
| LDL-C, mean (SD) | 2 | 121.6 (38.1) |
| ApoB, mean (SD) | 0 | 93.7 (25.2) |
| Lp(a), median [Q1,Q3] | 2 | 25.0 [15.4,61.9] |
| Triglycerides, median [Q1,Q3] | 0 | 77.0 [55.8,103.8] |
| hs-CRP, median [Q1,Q3] | 1 | 0.8 [0.4,1.4] |
| HbA1c, median [Q1,Q3] | 0 | 5.5 [5.2,5.6] |
| Glucose, median [Q1,Q3] | 0 | 93.0 [87.8,102.0] |
| Insulin, median [Q1,Q3] | 0 | 6.9 [5.2,10.3] |
| HOMA-IR, median [Q1,Q3] | 0 | 1.6 [1.2,2.4] |
| oxLDL, median [Q1,Q3] | 6 | 47.5 [29.5,92.8] |
| eGFR, mean (SD) | 0 | 88.6 (12.8) |

Continuous variables are mean (SD), except variables with absolute skewness above 1, which are median [Q1, Q3]. Categorical variables are n (%).

For evaluation of associations with both binary existence of “inflammation”, as defined as having an FAI-Score of 75 or higher, or with FAI-Score as a continuous variable, the primary prespecified hypothesis family consisted of ApoB, non-HDL-C, hs-CRP, HbA1c, age, and sex. All remaining metabolic, hemodynamic, treatment, and imaging variables were considered exploratory secondary variables. Relationships between selected biomarkers and clinical measures and maximum vessel FAI-Score percentile are shown in Figure 1.

**Figure 1.**
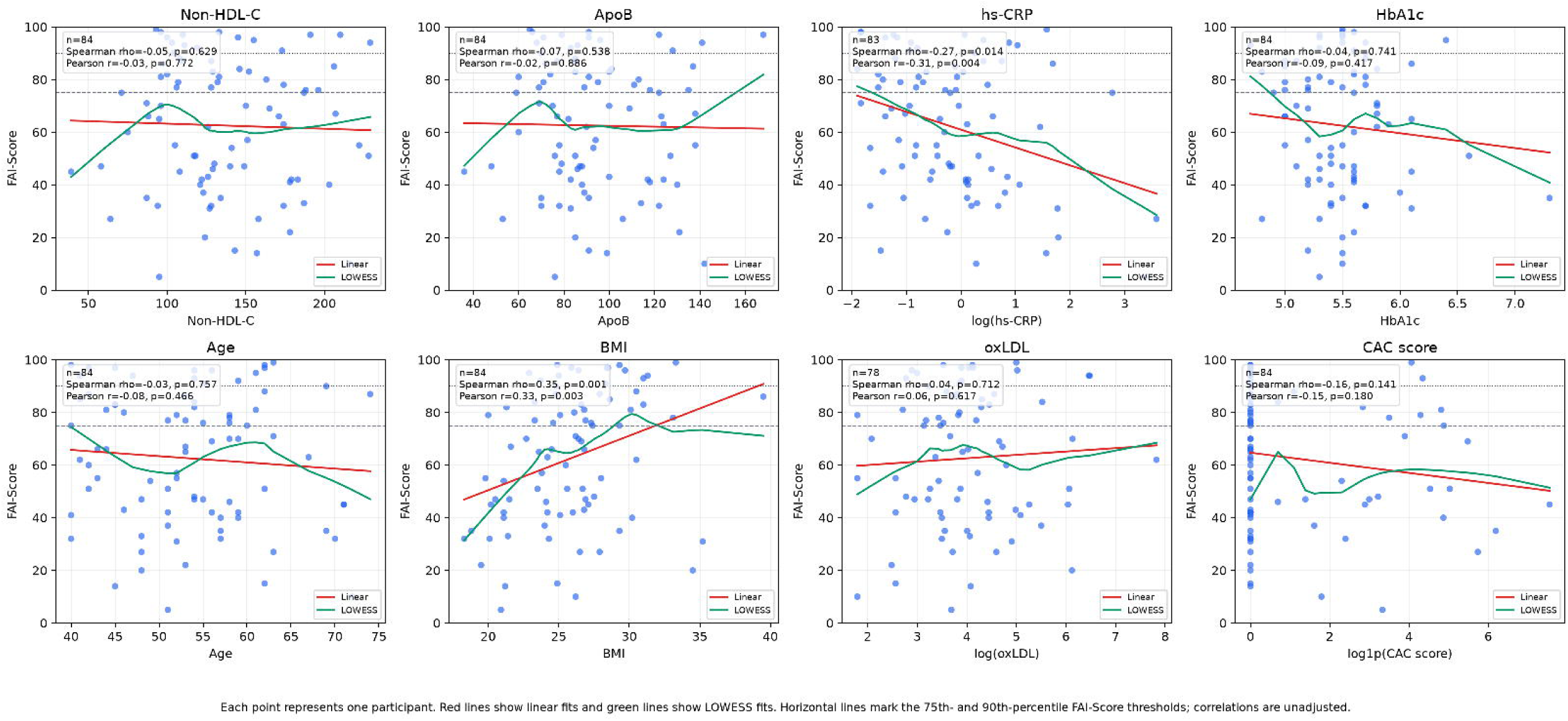
Continuous biomarkers and maximum vessel FAI-Score percentile

### Prevalence and Anatomical Extent of Perivascular Inflammation

Elevated FAI-Score was common in the cohort. Overall, 36 of 84 participants (42.9%; 95% CI, 32.8-53.5%) had an FAI-Score at or above the prespecified 75th-percentile threshold in at least one major epicardial coronary artery. The maximum vessel percentile was 75th–89th in 21 participants (25%; 95% CI,17.0%–35.2%) and ≥90th in 15 participants (17.9%; 95% CI, 11.1%–27.4%) (Table 3).

**Table 3.** FAI-Score distribution.

| Measure | Result |
| --- | --- |
| Maximum vessel FAI-Score percentile | 62.5 (25.2) |
| LAD FAI-Score percentile | 47.1 (27.3) |
| LCx FAI-Score percentile | 44.9 (27.6) |
| RCA FAI-Score percentile | 52.9 (27.3) |
| High FAI-Score, any vessel ≥75th percentile | 36/84 (42.9%; 95% CI 32.8%–53.5%) |
| Vessels ≥75th percentile: 0 | 48/84 (57.1%) |
| Vessels ≥75th percentile: 1 | 21/84 (25.0%) |
| Vessels ≥75th percentile: 2 | 8/84 (9.5%) |
| Vessels ≥75th percentile: 3 | 7/84 (8.3%) |
| LAD ≥75th percentile | 18/84 (21.4%; 95% CI 14.0%–31.3%) |
| LCx ≥75th percentile | 19/84 (22.6%; 95% CI 15.0%–32.6%) |
| RCA ≥75th percentile | 21/84 (25.0%; 95% CI 17.0%–35.2%) |
| Very high FAI-Score, any vessel ≥90th percentile | 15/84 (17.9%; 95% CI 11.1%–27.4%) |
| Vessels ≥90th percentile: 0 | 69/84 (82.1%) |
| Vessels ≥90th percentile: 1 | 10/84 (11.9%) |
| Vessels ≥90th percentile: 2 | 2/84 (2.4%) |
| Vessels ≥90th percentile: 3 | 3/84 (3.6%) |
| LAD ≥90th percentile | 8/84 (9.5%; 95% CI 4.9%–17.7%) |
| LCx ≥90th percentile | 4/84 (4.8%; 95% CI 1.9%–11.6%) |
| RCA ≥90th percentile | 11/84 (13.1%; 95% CI 7.5%–21.9%) |

Continuous variables are mean (SD), except variables with absolute skewness above 1, which are median [Q1, Q3]. Categorical variables are n (%).
Wilson score 95% confidence intervals without continuity correction are shown for FAI-Score prevalence estimates.

Among the 36 participants meeting the threshold, 21 had single-vessel involvement, eight had two-vessel involvement, and seven had three-vessel involvement (Table 3). The RCA most frequently exhibited values ≥90th percentile, occurring in 11 participants (13.1% of the overall cohort).

### Circulating Biomarkers and Clinical Risk in Relation to FAI-Score Elevation

A subset of biomarkers from standardized and specialized blood panels was chosen to evaluate against FAI-Score for correlative and predictive utility and are listed in Table 2. None of these, including ApoB, non-HDL cholesterol, HbA1c, hs-CRP, and age, had a statistically significant association with binary inflammation at either the 75th or 90th percentile. When compared with binary very high inflammation, there were similarly no associations.

A non-significant inverse association was shown between elevated hs-CRP and binary inflammation (rank-biserial r=-0.16, AUC=0.42, p=0.223, primary-family FDR q=0.666; Figure 2). In post-hoc analysis, two members presenting with extremely elevated hs-CRP most likely due to illness were removed from the cohort as potential outliers. The inverse relationship remained but was diminished, more closely approximating chance level discrimination (r=-0.12, AUC=0.44, p=0.355). When treating the FAI percentile as a continuous variable rather than a categorical, hs-CRP was also inversely correlated with FAI-Score (Spearman ρ=−0.27, p=0.014, primary-family q=0.072; Figure 1), but again the association weakened with the removal of the same two members (ρ=−0.22, p=0.053; q=0.265). In exploratory multivariable OLS analysis, natural-log transformed hs-CRP remained inversely correlated with FAI-Score (β=−9.51 percentile points per one-unit increase in ln[hs-CRP]; 95% CI −13.88 to −5.14, p < 0.001, adjusted R²=0.24).

**Figure 2.**
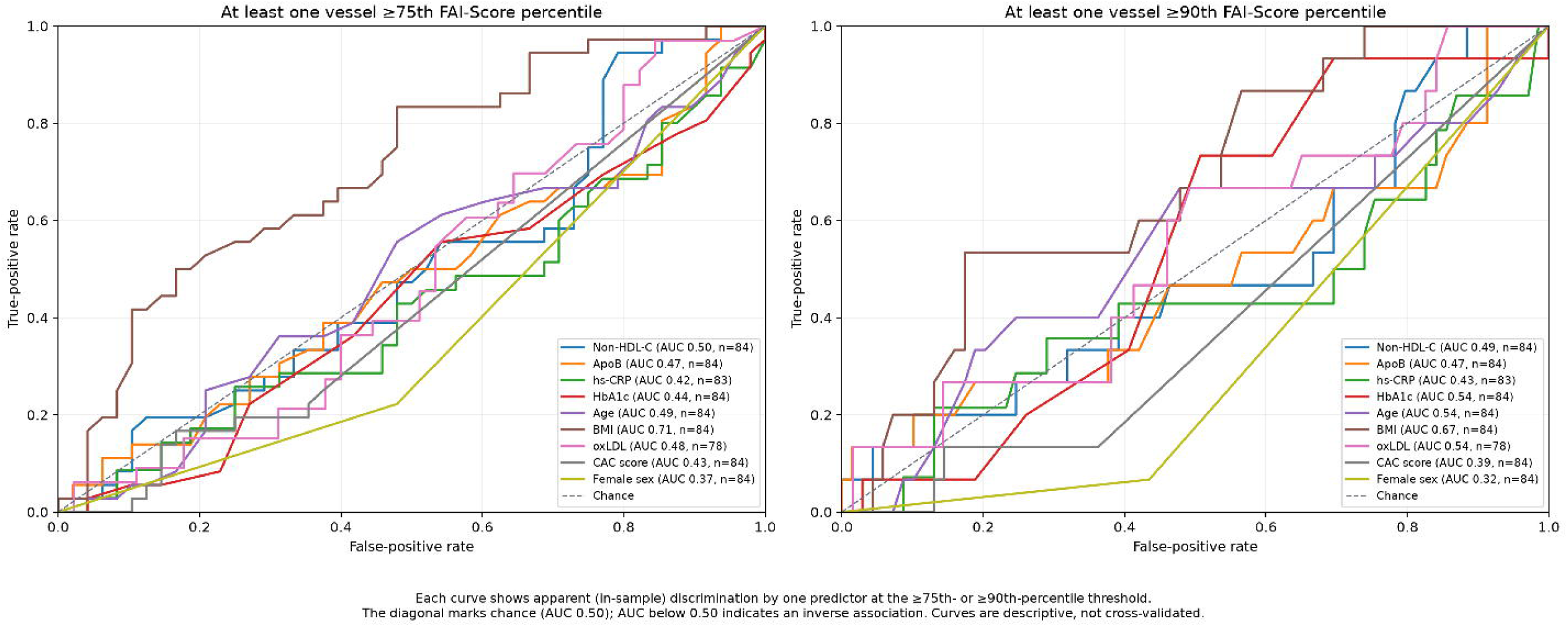
Exploratory univariate ROC curves for FAI-Score elevation

### Sex Differences in FAI-Score Elevation

Men had a higher prevalence of elevated FAI-Score than women at both thresholds. At least one vessel met the ≥75th-percentile threshold in 28 of 53 men (52.8%, Wilson 95% CI 39.7%-65.6%) and eight of 31 women (25.8%, CI 13.7%-43.2%), corresponding to an unadjusted odds ratio of 0.31 for women versus men (95% CI 0.1-0.89; p=0.016, primary-family FDR q=0.094). At the ≥90th-percentile threshold, the corresponding proportions were 14 of 53 men (26.4%) and one of 31 women (3.2%, OR 0.095, 95% CI 0.02-0.69; FDR q=0.08).

In post hoc logistic regression models including sex and BMI, the female sex association was weakened at the ≥75th-percentile threshold (adjusted OR 0.57, 95% CI 0.19-1.69, p=0.308). At the ≥90th-percentile the adjusted estimate remained inverse but imprecise (adjusted OR 0.11, 95% CI 0.013-1.004, p=0.0504).

### Exploratory Metabolic and Clinical Associations

Exploratory screening across 96 secondary association tests identified several nominal metabolic and hemodynamic associations, although only one met the significance threshold after false discovery rate correction across all tests.

Body mass index (BMI) showed the strongest observed correlation among the continuous secondary variables with person-level FAI-Score elevation and was the only secondary association to meet the all-test false discovery rate threshold. Median BMI was 27.1 kg/m² (IQR, 24.8–29.4) among participants with elevated FAI-Score versus 24.4 kg/m² (IQR, 21.4–26.7) among those below the threshold (rank biserial r=0.42, AUC=0.71, Mann-Whitney p=0.001, all-test FDR q=0.028).

Homeostatic Model Assessment of Insulin Resistance (HOMA-IR) and systolic blood pressure (SBP) displayed concordant, positive associations that did not reach significance. Median HOMA-IR was 2.0 (IQR 1.3-2.9) among inflamed participants vs 1.5 (IQR 1.1-2.0) among non-inflamed participants (r=0.24, AUC=0.62, p=0.057, q=0.190), whereas median SBP was 120.5 mmHg (IQR 110.0–130.0) versus 112.5 mmHg (IQR 106.8–125.3), respectively (r=0.24, AUC=0.62, p=0.061, q=0.190). Participants receiving antihypertensive therapy had higher unadjusted odds of FAI-Score elevation (9 of 13 [69.2%] vs. 25 of 71 [35.2%]; OR, 3.61,95% CI, 0.90–17.65, Pearson chi-square p=0.037; FDR q=0.186). Hypertension was similarly nominally associated (OR=3.26, p=0.040, q=0.186). Conversely, HDL cholesterol (r=−0.27, p=0.034, q=0.186) exhibited a weak nominal inverse association.

None of the six active smokers met the FAI-Score elevation threshold. Although the chi-squared test yielded p = 0.035, Fisher’s exact testing, used because of sparse cell counts, did not reach statistical significance (q=0.186).

### FAI-Score Elevation in Relation to Coronary Plaque and Stenosis

Elevated FAI-Score was observed among participants without detectable coronary stenosis. Of 65 participants without stenosis, 30 (46.2%) had at least one vessel at or above the 75th percentile, and 15 (23.1%) had a vessel at or above the 90th percentile. Among 19 participants with stenosis, six (31.6%) met the ≥75th-percentile threshold, and none met the greater than 90th-percentile threshold (Fisher’s exact p = 0.018 for the ≥90th-percentile comparison).

Among 53 participants without detectable calcified or noncalcified plaque, 25 (47.2%) met the greater than 75th-percentile threshold and 11 (20.8%) met the greater than 90th-percentile threshold. Plaque presence was not significantly associated with person-level FAI-Score elevation at the greater than 75th-percentile threshold (Pearson chi-squared p=0.296).

### Exploratory Logistic Regression Prediction of Elevated FAI-Score

Exploratory L2-regularized logistic regression models that used ApoB, natural-log-transformed hs-CRP, HbA1c, age, female sex, and BMI showed modest discrimination for binary inflammation (nested five-fold cross-validated AUC=0.71, n=83). The models were not able to discriminate for binary very high inflammation (AUC=0.52, n=83). Given the small sample size and internal validation, these models should be treated as investigative and preliminary.

## Discussion

This study characterizes the prevalence, vascular distribution, and clinical correlates of CCTA-derived FAI-Score in an asymptomatic adult population undergoing preventive evaluation in the United States. Three principal findings emerged with potential implications for early cardiovascular risk stratification. First, 42.9% of participants had an FAI-Score at or above the 75th percentile in at least one coronary artery, including 17.9% at or above the 90th percentile. Second, circulating atherogenic lipoproteins, HbA1c, hs-CRP, and continuous PREVENT-ASCVD risk estimates showed no statistically significant associations with the binary FAI-Score outcome, and the evaluated clinical and biomarker models demonstrated limited discrimination in internal validation. Third, elevated FAI-Score percentiles were observed in participants without detectable coronary plaque or stenosis; all 15 participants with values ≥90th percentile had no stenosis, and 11 had no detectable plaque. Collectively, these findings highlight coronary inflammation imaging as a promising, potentially distinct dimension of preventive cardiovascular evaluation, not already encapsulated by conventional coronary and inflammatory risk biomarkers and anatomical assessment, and support further investigation of its prognostic and clinical utility in asymptomatic populations.

### Circulating Biomarkers and Local Coronary Inflammation

Circulating atherogenic lipoproteins remain a cornerstone of cardiovascular risk assessment and preventive pharmacotherapy.[7] In this cohort, however, ApoB and non-HDL cholesterol showed no statistically significant associations with person-level FAI-Score elevation, and median ApoB concentrations were nearly identical between participants above and below the threshold (87.5 vs. 88.5 mg/dL). Similarly, hs-CRP was not significantly associated with the binary FAI-Score outcome. When analyzed continuously, FAI-Score demonstrated a slight inverse correlation.

These findings are consistent with the biological distinction between circulating risk markers and local tissue responses. ApoB-containing lipoproteins contribute to subendothelial lipid retention and atherogenesis, while endothelial dysfunction, oxidative modification, macrophage activity, and cytokine signaling shape the inflammatory response within the vessel wall.[13] CANTOS demonstrated that targeted inhibition of interleukin-1β can reduce recurrent cardiovascular events without lowering LDL cholesterol.[14] A pooled analysis of PROMINENT, REDUCE-IT, and STRENGTH likewise found that hs-CRP was more strongly associated with subsequent cardiovascular events and mortality than LDL cholesterol among statin-treated patients.[15] These studies establish the clinical relevance of inflammation alongside lipid-related risk; our findings extend this line of investigation into primary prevention, suggesting that circulating atherogenic lipoprotein levels alone may not fully capture the local coronary inflammatory phenotype reflected by FAI-Score.

Perivascular fat imaging offers a window into this local biology. Paracrine signaling between the coronary vessel wall and adjacent adipose tissue alters adipocyte differentiation and lipid accumulation, producing measurable changes in CT fat attenuation.[16] By contrast, hs-CRP is a hepatic acute-phase reactant that reflects systemic inflammatory signaling and is influenced by adiposity, intercurrent illness, and treatment.[23] These measures therefore capture different aspects of the inflammatory process: a low hs-CRP should not be interpreted as evidence of coronary inflammatory quiescence. The inverse hs-CRP association in our cohort requires replication and should not be interpreted as protective. Overall, our findings support coronary inflammation imaging as a potentially complementary dimension of preventive assessment, warranting prospective evaluation of its incremental prognostic value and ability to guide care.

### Coronary Inflammation in the Absence of Detectable Plaque or Stenosis

In our cohort, elevated FAI-Score percentiles were observed in 47.2% of participants without detectable coronary plaque and 46.2% of those without stenosis. Notably, all 15 participants with values at or above the 90th percentile had no detectable stenosis, and 11 had no detectable plaque. These observations highlight the potential for perivascular fat imaging to identify an inflammatory imaging phenotype even when conventional anatomical assessment shows no detectable coronary atherosclerosis.

These findings are biologically coherent with the role of inflammation throughout the development and progression of atherosclerosis.[13] Anatomical imaging characterizes plaque presence, extent, and luminal narrowing, whereas FAI-Score reflects changes in the adjacent perivascular adipose tissue associated with local vascular inflammatory signaling.[16] This distinction raises the possibility that inflammation imaging could provide an earlier window into coronary disease biology. However, the absence of CCTA-detectable plaque does not establish the absence of microscopic disease, and our cross-sectional observations cannot determine whether FAI-Score elevation precedes plaque development.

Prior outcome studies strengthen the rationale for investigating this phenotype. CRISP-CT linked elevated perivascular fat attenuation with cardiac mortality,[17] while ORFAN demonstrated independent prognostic associations between standardized FAI-Score and cardiac mortality and major adverse cardiovascular events, including among patients without obstructive CAD.[19] Our findings extend the descriptive evidence into an asymptomatic preventive cohort and support longitudinal evaluation of whether elevated FAI-Score in participants without detectable plaque predicts plaque progression or cardiovascular events beyond established risk measures.

### Clinical and Translational Implications for Primary Prevention

These findings highlight the potential to extract additional biological information from CCTA already obtained during preventive assessment. Elevated FAI-Score in participants without detectable plaque suggests an opportunity to refine coronary phenotyping beyond anatomical assessment alone. Whether this information meaningfully changes risk estimates or treatment decisions is a central question for prospective investigation. Identifying asymptomatic individuals with elevated FAI-Score creates an opportunity to investigate whether this imaging phenotype can help target preventive pharmacotherapy, including lipid-lowering and anti-inflammatory strategies. Prospective trials could determine whether FAI-Score identifies individuals who derive additional benefit from treatment beyond selection based on established risk factors.

Longitudinal therapeutic monitoring represents another promising application. Singh and colleagues demonstrated treatment-associated changes in left-main coronary FDG uptake after atorvastatin, supporting the responsiveness of coronary inflammatory imaging measures to therapy.[24] This provides a biological rationale for studying serial FAI-Score, but does not establish it as a surrogate for treatment benefit. Studies in asymptomatic populations should assess reproducibility, define changes that exceed measurement variability, and determine whether changes in FAI-Score track subsequent clinical outcomes.

Ultimately, prospective studies should establish whether FAI-guided care improves cardiovascular outcomes while evaluating downstream testing, cost-effectiveness, and the radiation and contrast exposure associated with any additional CCTA examinations.

### Study Limitations

Several limitations should inform interpretation and future investigation. The modest sample size and single, self-referred preventive-clinic population limit precision and generalizability. Multiple comparisons, small subgroups, and reliance on internal validation make the association and classification findings exploratory. Medication use and other unmeasured factors may have influenced both biomarkers and FAI-Score. FAI-Score is an imaging marker of inflammation, and the clinical meaning of the selected percentile thresholds in this population requires validation. Finally, the absence of longitudinal cardiovascular outcomes or an intervention comparison prevents conclusions about incremental prognosis, treatment benefit, or screening utility. The cross-sectional design also cannot establish that FAI-Score elevation precedes plaque formation. These data therefore do not yet support population-wide CCTA screening or treatment based solely on FAI-Score elevation.

### Conclusions

Elevated coronary FAI-Score percentiles were present in 42.9% of this asymptomatic preventive cohort, including among participants without detectable coronary plaque or luminal stenosis. Models incorporating routine clinical and biochemical measures, including ApoB, non-HDL-C, and hs-CRP, did not reliably predict FAI-Score elevation, and its prevalence did not differ significantly across the evaluated PREVENT-ASCVD risk categories. These findings highlight coronary inflammation imaging as a potentially distinct dimension of preventive cardiovascular assessment. If prospective studies establish incremental prognostic value and benefit from FAI-guided care, this approach could expand opportunities to identify and treat cardiovascular risk before a first clinical event.

## Data Availability

All relevant aggregate data supporting the conclusions of this study are contained within the manuscript and its supplementary material. Individual-level participant data, including raw imaging files and electronic health records, are not available for public distribution or sharing due to patient privacy protections and ethical governance constraints.

## Declaration

During the preparation of this manuscript, the authors utilized a large language model to assist with literature synthesis, initial text drafting, and language refinement across the Introduction, Materials & Methods, Results and Discussion sections. After utilizing these tools, the authors critically reviewed, verified, and substantively edited all generated content to ensure scientific rigor, clinical fidelity, and contextual accuracy against original primary sources. The authors confirm that the study design, IRB protocol execution, machine learning models, statistical analyses, and empirical data reported herein were conceived, executed, and validated independently by the investigators. The authors assume full accountability and responsibility for the integrity, veracity, and final content of this publication.

